# Effect of an intensive lifestyle intervention on circulating biomarkers of atrial fibrillation-related pathways

**DOI:** 10.1101/2023.04.28.23288131

**Authors:** Linzi Li, Alvaro Alonso, Dora Romaguera, Angel M. Alonso-Gómez, Cristina Razquin, Lucas Tojal-Sierra, Miquel Fiol, Miguel Angel Martínez-González, Vinita Subramanya, Jordi Salas-Salvadó, Montserrat Fito, Estefanía Toledo

## Abstract

Determining the effect of lifestyle interventions on blood concentrations of biomarkers of atrial fibrillation (AF)-related pathways could help understand AF pathophysiology and contribute to AF prevention. We studied 471 participants enrolled in the PREDIMED-Plus trial, a Spanish randomized trial in adults with metabolic syndrome. Eligible participants were randomized 1:1 to an intensive lifestyle intervention, emphasizing physical activity, weight loss, and adherence to an energy-reduced Mediterranean diet or to a control group. Serum biomarkers [carboxy-terminal propeptide of procollagen type I (PICP), high-sensitivity troponin T (hsTnT), high-sensitivity C reactive protein (hsCRP), 3-nitrotyrosine (3-NT), and N-terminal propeptide of B-type natriuretic peptide (NT-proBNP)] were measured at baseline, 3 and 5 years after randomization. Mixed models were used to evaluate the effect of intervention on changes in biomarkers through year 5. Mediation analysis was performed to examine the proportion mediated by each component of the intervention. Compared to the control group, participants in the intervention group experienced greater decreases in hsCRP (-16%, 95% confidence interval (CI) - 28%, -1%) or smaller increases in 3-NT (-15%, 95% CI -25%, -4%) and NT-proBNP (-13%, 95% CI - 25%, 0%). The intervention had minimal impact on hsTnT (-3%, 95% CI -8%, 2%) or PICP concentrations (-0%, 95% CI -9%, 9%). The effect of the intervention on hsCRP was primarily mediated by weight loss (73% and 66% at years 3 and 5). In conclusion, a dietary and lifestyle intervention for weight-loss favorably affected concentrations of hsCRP, 3-NT, and NT-proBNP, pointing to specific mechanisms in pathways linking lifestyles and AF.

**Condensed abstract:** In this study, we assessed the impact of an intensive lifestyle intervention on blood biomarkers related to atrial fibrillation (AF) pathways in 471 participants from the PREDIMED-Plus trial. The intervention focused on physical activity, weight loss, and an energy-reduced Mediterranean diet. Over five years, the intervention group showed greater decreases in high-sensitivity C reactive protein and smaller increases in 3-nitrotyrosine and N-terminal propeptide of B-type natriuretic peptide. Weight loss primarily mediated the effect of the intervention on high-sensitivity C reactive protein. These findings suggest that dietary and lifestyle changes may influence biomarkers involved in AF pathophysiology.

**Key points:** Randomized clinical trial, lifestyle, biomarker, atrial fibrillation

## Introduction

Atrial fibrillation (AF), a common sustained cardiac arrhythmia, has become a major public health problem with its growing impact on the population.^1^ In 2020, approximately 50 million people were affected by AF globally.^2, 3^ In the US, over 5 million people had AF in 2010, and it was expected to increase to 12.1 million in 2030.^2, 4^ The lifetime risk for AF has been estimated to be between 1 in 3 and 1 in 5.^5, 6^ Having AF increases the risk of many adverse outcomes, including ischemic stroke, heart failure (HF), hospitalization, and mortality, compounding the overall burden in public health.^5, 7, 8^ To date, effective detection and prevention strategies for AF are still lacking, partly due to an incomplete understanding of AF etiopathogenesis and pathophysiology. In animal models and AF patients, researchers have found evidence showing that specific electrical triggers act on a vulnerable structural and functional atrial substrate at the onset of AF.^9^ It has been suggested that atrial remodeling and fibrosis are significant features of the substrate facilitating AF onset. Previous studies have found several circulating biomarkers associated with increased risk of AF in AF-related pathways: carboxy-terminal propeptide of procollagen type I (PICP, marker of cardiac fibrosis), high-sensitivity troponin T (hsTnT, marker of myocardial damage), high-sensitivity C reactive protein (hsCRP, marker of inflammation), 3-nitrotyrosine (3-NT, marker of oxidative stress), and N-terminal propeptide of B-type natriuretic peptide (NT-proBNP, marker of atrial stretch). ^10–15^ These biomarkers characterize the atrial substrate and could help understand pathways of AF onset.

Lifestyles also influence the atrial substrate and AF risk. Obesity, diet, and physical activity may play a role in AF development. Observational studies have reported that overweight or obese people had an increased risk of AF, and weight gain was associated with higher AF risk.^16–18^ Our prior work in the PREDIMED trial found that a Mediterranean diet (MedDiet) enriched with extra-virgin olive oil (EVOO) reduced the risk of AF by 38%, compared to a control low-fat diet intervention.^19^ Additionally, the MedDiet intervention was beneficial in AF pathogenesis involved pathways by reducing inflammation, oxidative stress, and atrial stretch.^20–22^ The effects of physical activity on AF are inconsistent in different populations. In endurance athletes and middle-aged adults engaging in vigorous exercise, the risk of developing AF increased. ^23, 24^ In the elderly with moderate physical activity, the risk of AF was lower than those with no physical activity. ^17, 25^ It remains unclear how increased physical activity impacts the risk of AF and AF substrate in the general population.

Previous literature and our work in the PREDIMED trial suggested that lifestyle changes favorably affect AF substrate determinants (fibrosis, myocardial damage, inflammation, oxidative stress, and atrial stretch) and may lead to a decreased AF risk. Because the underlying pathophysiology of AF is multifactorial and complex,^26^ the effect of lifestyle changes on pathways related to atrial substrates development needs to be specified. Recognizing the role of lifestyle modification in AF development can inform preventive cardiology recommendations and public health policymaking. Therefore, in a new randomized trial, PREDIMED-Plus, we hypothesized that an intensive lifestyle intervention (ILI) program emphasizing physical activity, weight loss, and adherence to an energy-reduced MedDiet (erMedDiet) would lead to favorable changes in the circulating concentrations of PICP (cardiac fibrosis), hsTnT (myocardial damage), hsCRP (inflammation), 3-NT (oxidative stress), and NT-proBNP (atrial stretch). We aimed to 1) assess the effect of ILI on the concentrations of blood biomarkers related to AF pathways, and 2) evaluate the mediation proportion that each intervention component contributed to the relationship between ILI and biomarkers’ concentrations.

## Methods

### Study design and population

The PREDIMED-Plus study is a multicenter randomized trial in overweight and obese adults focusing on the primary prevention of cardiovascular disease (CVD). Between 2013 and 2016, 3,574 men (age 55-75) and 3,300 women (age 60-75) who had a body mass index (BMI) ≥ 27 and < 40 kg/m^2^ and met at least three components of the metabolic syndrome were recruited from 23 study sites in Spain. Exclusion criteria included documented history of previous CVD (including uncontrolled AF) at enrollment, and active malignant cancer or history of malignancy within five years.^27^ Upon enrollment, 6874 participants were randomized 1:1 to an ILI program or a control group.^27^ Computer-generated random allocation was centrally done in blocks of six participants and stratified by sex, age (<65, 65–70, >70 years) and center. The randomization procedure was internet-based and blinded to all staff and to the principal investigators of each center. Spouses of participants who wished to belong to the same group were randomized together, and we used the couple as unit of randomization. In the specific cases of couples in which the first spouse was previously recruited at a different time, the last spouse entering the study was directly assigned (not randomized) to the same study arm than his/her partner.

ILI program is based on an erMedDiet, increased physical activity, and cognitive-behavior weight management, and the control intervention consists in low-intensity dietary advice on a MedDiet which is not reduced in calories (ad libitum total energy intake). The intervention was completed at the beginning of 2023, but the trial is ongoing, with a 2-year period of extended observation, without intervention until December 2024. Baseline and follow-up examinations were performed at randomization, at six months and year 1 post-randomization, and then annually among all the participants. As part of an ancillary study to the PREDIMED-Plus trial, echocardiographic studies were conducted in 3 PREDIMED-Plus study sites (University of Navarra, Araba University Hospital, Son Espases University Hospital; n = 566), and blood biomarkers were measured at baseline and years 3 and 5 after randomization in a sub-sample of those who had echocardiographic assessment.^28^

For this analysis, participants who completed baseline and at least one follow-up exam were included; subjects who had AF at baseline and lacked blood biomarker measurements at baseline and at both the year 3 and year 5 follow-up visits were excluded from this study (Figure 1). Institutional Review Boards at all participating institutions approved the study. Participants provided written informed consent at each visit.

**Figure 1.**
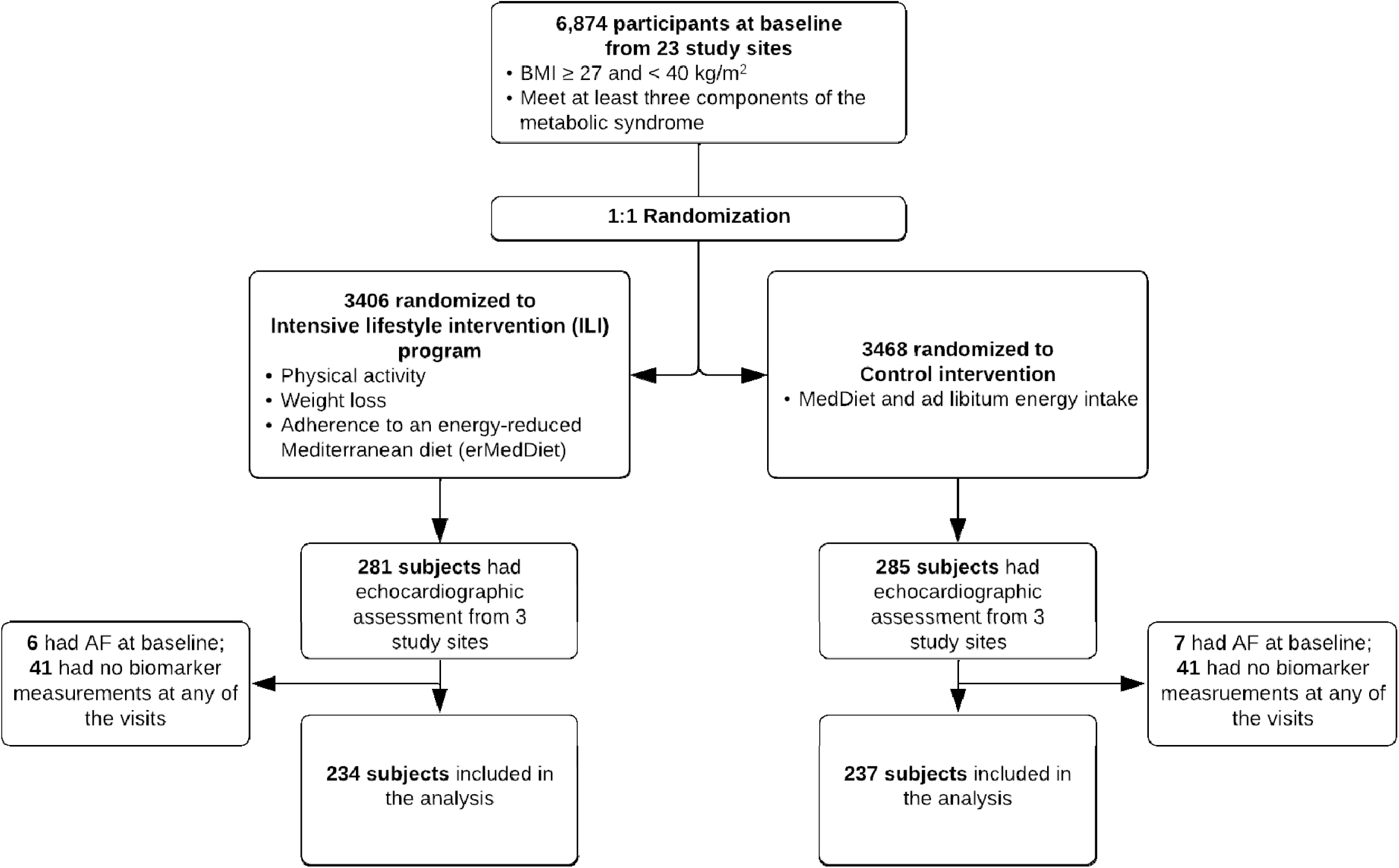
CONSORT chart of the study sample, PREDIMED-Plus trial

The trial was registered in 2014 at [www.isrctn.com/ISRCTN89898870]. The full protocol is available at www. https://www.predimedplus.com/en/project/.

### Intervention components and outcome measures

The ILI components were MedDiet adherence, physical activity (moderate to vigorous physical activity [MVPA], total physical activity [TPA]), and weight loss, defined using the 17-item erMedDiet questionnaire, MET-min/week from abbreviated Minnesota Leisure Time Physical Activity Questionnaire, and weight in kilograms, at baseline and each follow-up visits at years 3 and 5.^27^

The primary outcomes were five blood biomarker concentrations measured in serum samples at baseline and two follow-up exams: PICP (fibrosis), hsTnT (myocardial damage), hsCRP (inflammation), 3-NT (oxidative stress), and NT-proBNP (atrial stretch). HsTnT and NT-proBNP were measured by electrochemiluminescence immunoassay (ECLIA) and hsCRP by immunoturbidimetry on a Cobas 8000 autoanalyzer (Roche Diagnostics). Enzyme-linked immunosorbent assay technique (ELISA) was performed to measure 3-NT and PICP using commercially available assays. 3-NT was measured using the Human Nitrotyrosin ELISA kit (Abcam, Cambridge, UK) and PICP with MicroVue PICP EIA (Quidel, San Diego, CA, USA). All analyses were conducted blindly.

### Other covariates

In questionnaires at baseline, participants’ demographic characteristics (age, sex, origin, education, marital status), behavioral characteristics (smoking status, alcohol consumption), clinical measurements (BMI, systolic blood pressure [SBP], diastolic blood pressure [DBP], low-density lipoprotein cholesterol [LDLc], high-density lipoprotein cholesterol [HDLc], triglycerides, total cholesterol [TC]), medical history (diabetes), and medications (anti-hypertension, lipid-lowing medication) were collected. At baseline, years 3 and 5, fasting blood samples were collected. Using the Jaffe method, serum creatinine was measured by enzymatic reaction. The estimated glomerular filtration rate (eGFR) was calculated based on 2021 CKD-EPI creatinine equation.^29^

### Sample size estimation

Based on a 1:1 parallel design, with repeated measures at years 0, 3, and 5, power=0.9, 2-tailed alpha=0.005 (to account for the multiple echocardiographic variables and biomarkers considered in the substudy), a within-person correlation (rho) of 0.1 in the outcome variables, and a conservative sample size of 430, we would be able to detect a between-group difference in the slope of the annual rate of change of 0.105 standard deviations. Assumptions about differences in annual rate change were reasonable given the 1-year difference in change of C-reactive protein was 2.6 mg/dL (0.5 SD) in PREDIMED.^30^

### Statistical analysis

Baseline characteristics of the ILI and control arm were compared to examine if the randomization had balanced important covariates. We log transformed biomarker concentrations and these log-transformed values were used in all analyses. Mixed models were used to estimate the effect of ILI on changes in log-transformed biomarkers through years 3 and 5 with an intention-to treat approach. Follow-up time was modeled as a categorical variable (year 3 vs. year 0, year 5 vs. year 0). The model included random effects for the individual and for the family unit, since 24 participants had a single household cohabitant participating in the study. Effect of the intervention on biomarker change was estimated by including interaction terms between year and intervention. Furthermore, we conducted a mediation analysis to decompose the total effect in direct effect of the intervention and indirect effect mediated through any of the three ILI components (weight loss, adherence to the MedDiet, and physical activity), and then we calculated the proportion mediated, defined as the indirect effect divided by the total effect. The analysis was performed using the methods and SAS macro developed by Valeri and VanderWeele.^31^

In a stratified analysis, we ran mixed models in populations stratified by sex and age 65 years to examine ILI’s effect on biomarkers change. To minimize the impact of missing data of those who only came for one follow-up visit, we used multiple imputation by chained equations (MICE) to impute missing biomarker measurements and then ran the mixed models in 25 imputed datasets. Furthermore, we included age, sex, marital status, education level, smoking, and covariates that were not comparable between two arms at baseline in the mixed models to reduce potential confounding due to random imbalance. Lastly, to account for the impact of kidney function on the ability of how the biomarkers characterized the AF substrates, we adjusted for eGFR in all models. The results were presented in percent changes, calculated as [exp(coefficient with mixed model)-1]*100, along with 95% confidence intervals (CIs). CIs that do not cross the null can be considered statistically significant at the 0.05 level. SAS statistical software (v.9.4, SAS Institute Inc.) was used to complete all the analyses.

## Results

Four hundred and seventy-one participants were included in the analyses (Figure 1), with a mean age of 65 (standard deviation [SD] 4.9) years and BMI of 32 (SD 3.3) kg/m^2^. Of those, 193 (41%) were female, and 234 (50%) were assigned to the ILI arm. At baseline, participants in the ILI arm had a lower amount of MVPA (1792 vs. 2131 MET-min/week) and TPA (2400 vs. 2848 MET-min/week) and higher MedDiet score (8.0 vs. 7.8). Table 1 shows the baseline characteristics of the two arms.

**Table 1.** Baseline characteristics of participants by arms in PREDIMED-Plus trial.

|  | Total (n = 471) | ILI arm (n = 234) | Control arm (n = 237) |
| --- | --- | --- | --- |
| Age, yr | 65.1 (4.9) | 64.6 (5.0) | 65.6 (4.8) |
| Female, % | 193 (41.0) | 89 (38.0) | 104 (43.9) |
| Origin, % |  |  |  |
| European | 457 (97.0) | 227 (97.0) | 230 (97.1) |
| Latin American | 14 (3.0) | 7 (3.0) | 7 (3.0) |
| Marital status, % |  |  |  |
| Single | 27 (5.7) | 13 (5.6) | 14 (5.9) |
| Married/cohabiting | 372 (79.2) | 180 (76.9) | 192 (81.4) |
| Divorced | 44 (9.4) | 28 (12.0) | 16 (6.8) |
| Widower | 27 (5.7) | 13 (5.6) | 14 (5.9) |
| Education, % |  |  |  |
| Less than high school | 227 (48.2) | 106 (45.3) | 121 (51.1) |
| High school | 144 (30.6) | 80 (34.2) | 64 (27.0) |
| At least some college | 100 (21.2) | 48 (20.5) | 52 (21.9) |
| BMI, kg/m<sup>2</sup> | 32.2 (3.3) | 32.6 (3.4) | 31.9 (3.1) |
| SBP, mmHg | 141.5 (17.0) | 142.2 (18.2) | 140.7 (15.8) |
| DBP, mmHg | 79.1 (10.3) | 79.3 (10.6) | 79.0 (10.0) |
| Smoking, % |  |  |  |
| Never | 180 (38.2) | 89 (38.0) | 91 (38.4) |
| Former | 241 (51.2) | 118 (50.4) | 123 (51.9) |
| Current | 48 (10.2) | 27 (11.5) | 21 (8.9) |
| Alcohol consumption, g/day | 14.5 (18.9) | 14.6 (19.2) | 14.3 (18.7) |
| MedDiet adherence score | 7.9 (2.9) | 8.0 (2.8) | 7.8 (3.0) |
| MVPA, METs-min/week | 1962.7 (2230.9) | 1792.3 (2246.7) | 2130.9 (2207.1) |
| TPA, METs-min/week | 2625.5 (2280.3) | 2400.3 (2317.1) | 2847.8 (2226.0) |
| Diabetes, % | 146 (31.0) | 70 (29.9) | 76 (32.1) |
| Anti-hypertension medication, % | 362 (76.9) | 181 (77.4) | 181 (76.4) |
| Lipid-lowering medication, % | 243 (51.6) | 120 (51.3) | 123 (51.9) |
| Total cholesterol, mg/dL | 198.7 (35.9) | 197.8 (34.0) | 199.6 (37.7) |
| Triglycerides, mg/dL | 153.0 (71.0) | 151.5 (68.7) | 154.5 (73.4) |
| HDLc, mg/dL | 45.3 (10.8) | 45.5 (10.9) | 45.1 (10.7) |
| LDLc, mg/dL | 123.3 (31.3) | 122.2 (29.5) | 124.3 (32.9) |
| eGFR, mL/min/1.73 m<sup>2</sup> | 90.5 (12.3) | 91.6 (11.2) | 89.3 (13.2) |
| PICP, mg/mL | 99.0 (43.2) | 96.5 (38.4) | 101.6 (47.5) |
| HsTnT, ng/L | 9.4 (4.8) | 9.4 (5.4) | 9.3 (4.3) |
| hsCRP, mg/dL | 0.4 (0.5) | 0.4 (0.5) | 0.4 (0.5) |
| 3-NT, nM | 802.1 (705.4) | 848.4 (694.0) | 756.0 (715.0) |
| NT-proBNP, pg/mL | 79.2 (134.8) | 75.4 (88.9) | 82.9 (168.6) |

Notes: Data are shown as frequency (percentage) or mean (SD) for continuous variables of the sample. SBP: systolic blood pressure; DBP, diastolic blood pressure; MedDiet: Mediterranean Diet; HDLc: high-density lipoprotein cholesterol; LDLc: low-density lipoprotein cholesterol; MVPA: moderate to vigorous physical activity; TPA: total physical activity; eGFR: estimated glomerular filtration rate; PICP: C-terminal propeptide of procollagen type I; hsTnT: high sensitivity troponin T; hsCRP: high-sensitivity C reactive protein; 3-NT: 3-nitrotyrosine; NT-proBNP: N-terminal propeptide of B-type natriuretic peptide.

At years 3 and 5, 444 (94%) and 434 (92%) had available biomarker measurements. Over five years of follow-up, hsTnT, 3-NT, and NT-proBNP concentrations kept increasing in both ILI and control arms; PICP concentration increased in the control arm and first dropped at year 3 and then elevated in the ILI arm; hsCRP concentration decreased in the ILI arm and fluctuated in the control arm (Figure 2). The mean changes in log-transformed biomarkers concentrations were -0.02 for PICP, 0.11 for hsTnT, -0.08 for hsCRP, 0.09 for 3-NT, and 0.18 for NT-proBNP at year 3, and were -0.03 for PICP, 0.19 for hsTnT, - 0.15 for hsCRP, 0.12 for 3-NT, and 0.30 for NT-proBNP at year 5 compared to baseline. After five years, participants in the ILI arm had a smaller increase in 3-NT and NT-proBNP concentrations and a larger decrease in hsCRP concentration compared to participants in the control arm. The difference in percent changes of biomarker concentrations by intervention groups were -15% (95% CI -25%, -4%) for 3-NT, - 13% (95% CI -25%, 0%) for NT-proBNP, and -16% (95% CI -28%, -1%) for hsCRP. No significant changes in hsTnT and PICP concentration were observed between the two arms (-3%, 95% CI -8%, 2% for hs-TnT; 0%, 95% CI -9%, 9% for PICP) (Table 2).

**Figure 2.**
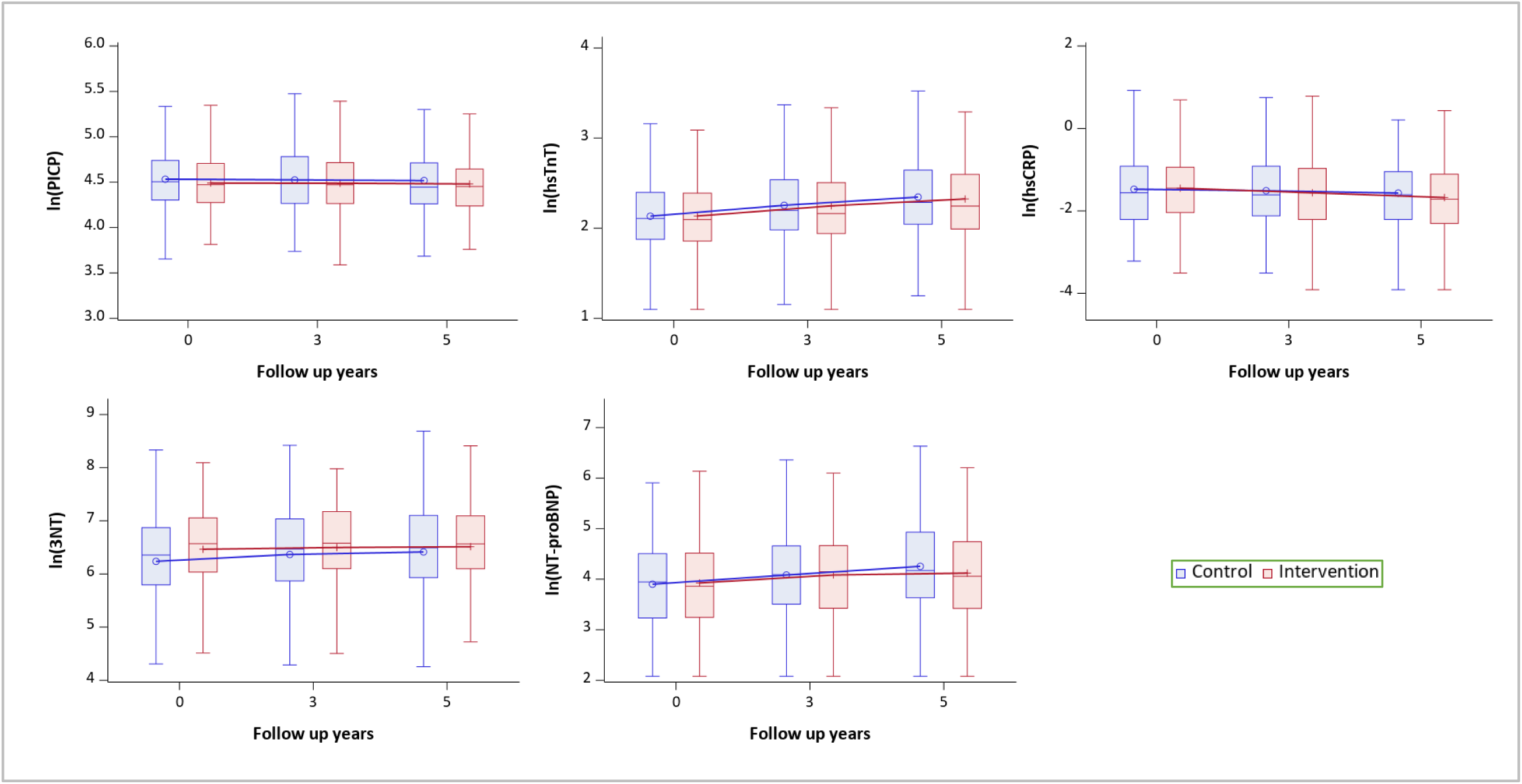
Log-transformed biomarker concentrations at baseline, years 3 and 5 Notes: Boxplots show minimum (Q1-1.5*IQR), Q1, median, Q3, maximum (Q3-1.5*IQR) of each log-transformed biomarker concentrations. Boxes at baseline, year 3 and year 5 are connected with a single line for the mean.

**Table 2.** Effect of an intensive lifestyle intervention (ILI) on selected circulating biomarkers, PREDIMED-PLUS trial.

|  | Control arm |  | ILI arm |  | Difference ILI - control |  |
| --- | --- | --- | --- | --- | --- | --- |
|  | Y3 vs baseline | Y5 vs baseline | Y3 vs baseline | Y5 vs baseline | Y3 vs baseline | Y5 vs baseline |
| PICP | -1% (-7%, 6%) | -2% (-8%, 5%) | -2% (-7%, 3%) | -3% (-8%, 2%) | -1% (-9%, 8%) | 0% (-9%, 9%) |
| Hs-TnT | 13% (9%, 17%) | 24% (19%, 28%) | 10% (6%, 15%) | 20% (15%, 24%) | -2% (-7%, 3%) | -3% (-8%, 2%) |
| HsCRP | -2% (-13%, 10%) | -7% (-17%, 5%) | -13% (-22%, -3%) | -21% (-30%, -12%) | -11% (-24%, 5%) | -16% (-28%, -1%) |
| 3-NT | 15% (4%, 26%) | 21% (10%, 33%) | 4% (-4%, 12%) | 3% (-5%, 12%) | -10% (-20%, 2%) | -15% (-25%, -4%) |
| NT-proBNP | 22% (11%, 35%) | 44% (30%, 59%) | 17% (5%, 30%) | 25% (12%, 39%) | -4% (-17%, 11%) | -13% (-25%, 0%) |

Notes: Mixed models were used. Results are shown as estimate (95% confidence interval). All the concentrations of biomarkers were log-transformed. Percent changes in concentrations of biomarkers by arms and comparing intervention and control arm were presented, calculated as $[\exp(\text{coefficient})-1]*100$ .

### Mediation analysis

The effect of ILI on the change of hsCRP concentration over time was primarily mediated by weight loss (73% at year 3 and 66% at year 5). The natural indirect effects of ILI mediated through weight loss were - 10% at year 3 (95% CI -17%, -2%) and -11% at year 5 (95% CI -17%, -4%), while the total effects of ILI were -13% (95% CI -26%, 2%) and -16% (95% CI -29%, -1%). Weight loss did not mediate the effect of ILI on other biomarkers to a substantial amount. Similarly, we did not find evidence of mediation of ILI on biomarkers through MVPA, TPA, and MedDiet score (Table 3).

**Table 3.**
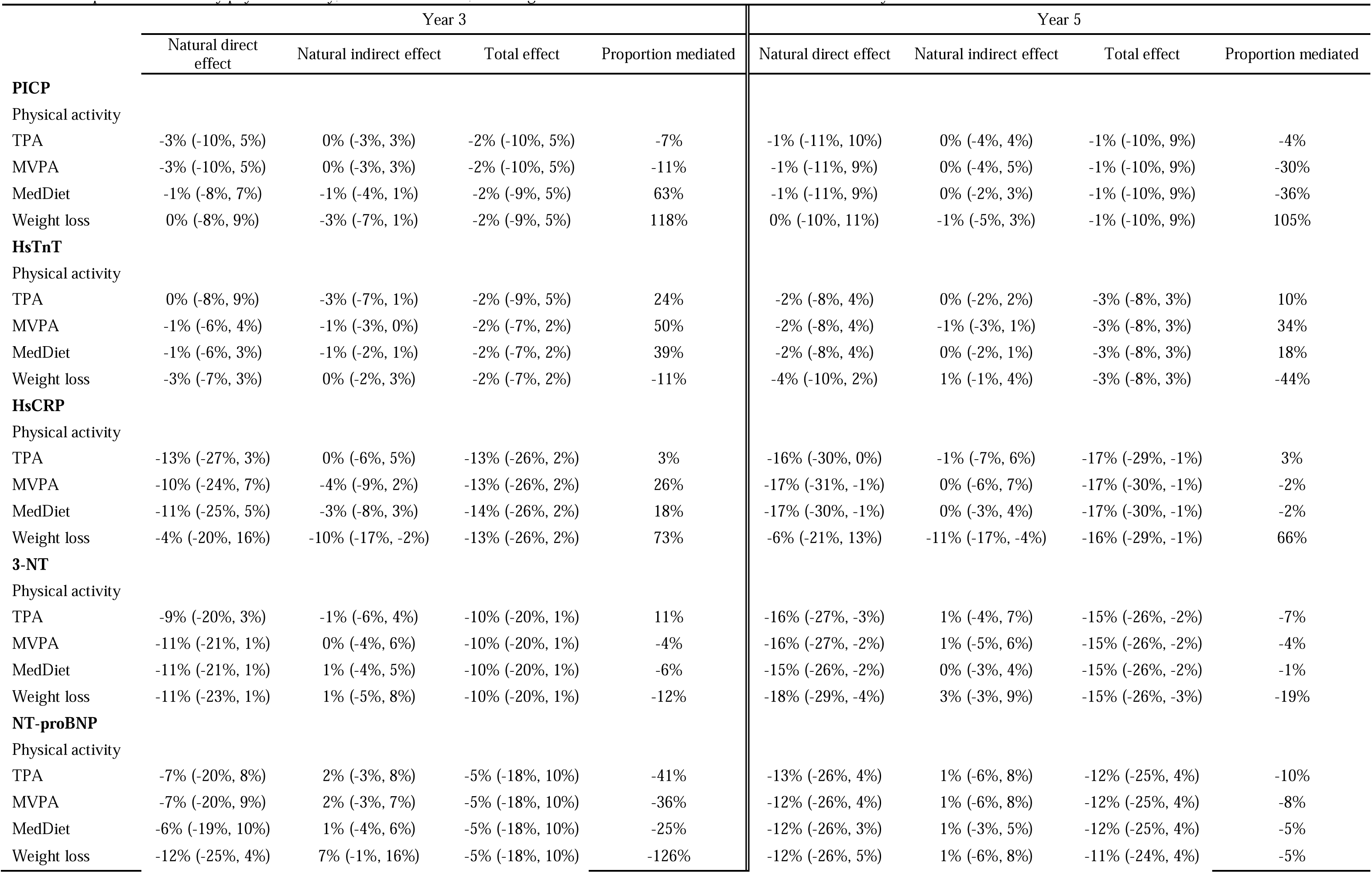
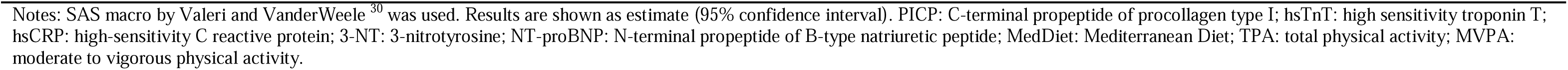
Proportion mediated by physical activity, Mediterranean diet, and weight loss in the effect of ILI on biomarkers over 3 or 5 years.

|  | Year 3 |  |  |  | Year 5 |  |  |  |
| --- | --- | --- | --- | --- | --- | --- | --- | --- |
|  | Natural direct effect | Natural indirect effect | Total effect | Proportion mediated | Natural direct effect | Natural indirect effect | Total effect | Proportion mediated |
| **PICP** |  |  |  |  |  |  |  |  |
| Physical activity |  |  |  |  |  |  |  |  |
| TPA | -3% (-10%, 5%) | 0% (-3%, 3%) | -2% (-10%, 5%) | -7% | -1% (-11%, 10%) | 0% (-4%, 4%) | -1% (-10%, 9%) | -4% |
| MVPA | -3% (-10%, 5%) | 0% (-3%, 3%) | -2% (-10%, 5%) | -11% | -1% (-11%, 9%) | 0% (-4%, 5%) | -1% (-10%, 9%) | -30% |
| MedDiet | -1% (-8%, 7%) | -1% (-4%, 1%) | -2% (-9%, 5%) | 63% | -1% (-11%, 9%) | 0% (-2%, 3%) | -1% (-10%, 9%) | -36% |
| Weight loss | 0% (-8%, 9%) | -3% (-7%, 1%) | -2% (-9%, 5%) | 118% | 0% (-10%, 11%) | -1% (-5%, 3%) | -1% (-10%, 9%) | 105% |
| **HsTnT** |  |  |  |  |  |  |  |  |
| Physical activity |  |  |  |  |  |  |  |  |
| TPA | 0% (-8%, 9%) | -3% (-7%, 1%) | -2% (-9%, 5%) | 24% | -2% (-8%, 4%) | 0% (-2%, 2%) | -3% (-8%, 3%) | 10% |
| MVPA | -1% (-6%, 4%) | -1% (-3%, 0%) | -2% (-7%, 2%) | 50% | -2% (-8%, 4%) | -1% (-3%, 1%) | -3% (-8%, 3%) | 34% |
| MedDiet | -1% (-6%, 3%) | -1% (-2%, 1%) | -2% (-7%, 2%) | 39% | -2% (-8%, 4%) | 0% (-2%, 1%) | -3% (-8%, 3%) | 18% |
| Weight loss | -3% (-7%, 3%) | 0% (-2%, 3%) | -2% (-7%, 2%) | -11% | -4% (-10%, 2%) | 1% (-1%, 4%) | -3% (-8%, 3%) | -44% |
| **HsCRP** |  |  |  |  |  |  |  |  |
| Physical activity |  |  |  |  |  |  |  |  |
| TPA | -13% (-27%, 3%) | 0% (-6%, 5%) | -13% (-26%, 2%) | 3% | -16% (-30%, 0%) | -1% (-7%, 6%) | -17% (-29%, -1%) | 3% |
| MVPA | -10% (-24%, 7%) | -4% (-9%, 2%) | -13% (-26%, 2%) | 26% | -17% (-31%, -1%) | 0% (-6%, 7%) | -17% (-30%, -1%) | -2% |
| MedDiet | -11% (-25%, 5%) | -3% (-8%, 3%) | -14% (-26%, 2%) | 18% | -17% (-30%, -1%) | 0% (-3%, 4%) | -17% (-30%, -1%) | -2% |
| Weight loss | -4% (-20%, 16%) | -10% (-17%, -2%) | -13% (-26%, 2%) | 73% | -6% (-21%, 13%) | -11% (-17%, -4%) | -16% (-29%, -1%) | 66% |
| **3-NT** |  |  |  |  |  |  |  |  |
| Physical activity |  |  |  |  |  |  |  |  |
| TPA | -9% (-20%, 3%) | -1% (-6%, 4%) | -10% (-20%, 1%) | 11% | -16% (-27%, -3%) | 1% (-4%, 7%) | -15% (-26%, -2%) | -7% |
| MVPA | -11% (-21%, 1%) | 0% (-4%, 6%) | -10% (-20%, 1%) | -4% | -16% (-27%, -2%) | 1% (-5%, 6%) | -15% (-26%, -2%) | -4% |
| MedDiet | -11% (-21%, 1%) | 1% (-4%, 5%) | -10% (-20%, 1%) | -6% | -15% (-26%, -2%) | 0% (-3%, 4%) | -15% (-26%, -2%) | -1% |
| Weight loss | -11% (-23%, 1%) | 1% (-5%, 8%) | -10% (-20%, 1%) | -12% | -18% (-29%, -4%) | 3% (-3%, 9%) | -15% (-26%, -3%) | -19% |
| **NT-proBNP** |  |  |  |  |  |  |  |  |
| Physical activity |  |  |  |  |  |  |  |  |
| TPA | -7% (-20%, 8%) | 2% (-3%, 8%) | -5% (-18%, 10%) | -41% | -13% (-26%, 4%) | 1% (-6%, 8%) | -12% (-25%, 4%) | -10% |
| MVPA | -7% (-20%, 9%) | 2% (-3%, 7%) | -5% (-18%, 10%) | -36% | -12% (-26%, 4%) | 1% (-6%, 8%) | -12% (-25%, 4%) | -8% |
| MedDiet | -6% (-19%, 10%) | 1% (-4%, 6%) | -5% (-18%, 10%) | -25% | -12% (-26%, 3%) | 1% (-3%, 5%) | -12% (-25%, 4%) | -5% |
| Weight loss | -12% (-25%, 4%) | 7% (-1%, 16%) | -5% (-18%, 10%) | -126% | -12% (-26%, 5%) | 1% (-6%, 8%) | -11% (-24%, 4%) | -5% |

Notes: SAS macro by Valeri and VanderWeele<sup>30</sup> was used. Results are shown as estimate (95% confidence interval). PICP: C-terminal propeptide of procollagen type I; hsTnT: high sensitivity troponin T; hsCRP: high-sensitivity C reactive protein; 3-NT: 3-nitrotyrosine; NT-proBNP: N-terminal propeptide of B-type natriuretic peptide; MedDiet: Mediterranean Diet; TPA: total physical activity; MVPA: moderate to vigorous physical activity.

### Stratified analysis

Sex slightly modified the effect of the intervention on some of the biomarkers without significant interaction. Among females, the ILI did not affect hsCRP concentration (difference of change between ILI and control at year 5: 5%, 95% CI -15%, 30%), while that difference was remarkably higher among males (difference: -27%, 95% CI -42%, -8%) (p-value for interaction: 0.31) (Supplemental Table 1). Similarly, the difference in NT-proBNP concentration change between the two arms was larger among males. Females had a higher difference of 3-NT concentration change between two arms than males. The results of PICP and hsTnT among males and females were comparable with those in the overall population. Participants aged < 65 years had a higher change in hsCRP concentration and a lower change in 3-NT concentration in the ILI arm than the control arm compared to participants aged ≥ 65 years. Those aged <65 years and ≥ 65 years had similar results for PICP, hsTnT, and NT-proBNP. None of the interactions between age ≥ 65 years and ILI on the changes of biomarker concentrations were significant.

### Sensitivity analysis

After imputing the missing biomarker concentrations, the results were consistent with those in the primary analyses (Supplemental Table 2). Adjusting for potential confounders (sex, age, education level, marital status, smoking, Mediterranean diet adherence score, and TPA) at baseline also did not notably change the results (Supplemental Table 3), as participants in the ILI arm experienced a smaller increase in 3-NT and NT-proBNP concentration and a greater decrease in hsCRP concentration. Also, considering kidney function at baseline, years 3 and 5 in the models did not result in any substantial change in the effect of ILI on the concentrations of biomarkers.

## Discussion

In this study, we found that over five years of follow-up, there were increases in hsTnT, 3-NT, and NT-proBNP concentrations and a decrease in hsCRP concentration in the total study population. Compared to the control arm, the ILI arm showed lower increases in 3-NT and NT-proBNP, and a higher decrease in hsCRP. Additionally, the total effect of ILI on the change of hsCRP concentration was noticeably mediated by weight loss at both years 3 and 5, while no evidence was found of physical activity and erMedDiet adherence mediating any of the effects of ILI on biomarkers.

Our results suggested that ILI has a stronger impact than the control program on concentrations of selected biomarkers that can characterize the atrial substrate and AF-related pathways, including inflammation (hsCRP), atrial stretch (NT-proBNP), and oxidative stress (3-NT). Weight-loss targeted intensive lifestyle intervention can effectively reduce AF risk through these pathways. A preliminary analysis of the PREDIMED-Plus study has reported that ILI resulted in improvements in weight loss, diet adherence, and other CVD risk factors.^32^ Our study results were fairly consistent with published studies investigating weight loss and biomarker levels. Previous experimental studies have documented that overweight and obesity are associated with an increased serum hsCRP, and weight loss could decrease serum hsCRP. In the POUNDS LOST study, a randomized weight-loss trial of 811 adults, it was found that hsCRP levels were decreased by a similar magnitude in different weight-loss treatment arms.^33^ An interventional study of 59 obese females who received a 6-month dietary therapy in Spain revealed that weight loss was associated with reduced inflammatory response and improved oxidative stress, with a lower level of serum hsCRP.^34^ Another randomized clinical trial of 243 menopausal women in the Netherlands also showed that hsCRP level was reduced by a modest amount of weight loss.^35^ Among 206 children aged 10-18 years in Germany, exercise and diet targeting weight loss decreased hsCRP concentration. ^36^ These studies implied that intentional weight loss could be an effective intervention for alleviating inflammation and potentially oxidative stress among various populations. Some studies reported a relationship between weight loss and NT-proBNP, either short- or long-term. ^37–40^ A nested case-control study has found that the 3-NT level was significantly higher among patients with metabolic syndrome and resultant cardiovascular autonomic neuropathy than the controls, and a 24-week lifestyle intervention highly reduced 3-NT concentration among the patients. ^41^ Similar reduction in 3-NT concentration by 6-week lifestyle intervention on diet and physical activity was reported among 19 overweight or obese adults in Canada. ^42^ There were inconsistent results regarding the effect of lifestyle intervention on high-sensitivity troponin I and T. ^43–45^ PICP level was found higher among HF patients with coexisting AF than patients with sinus rhythm. ^46^ However, little is known about how lifestyle changes influence PICP.

Prior investigations in US general populations recorded that changes in hsTnT and NT-proBNP concentrations were directly related to the risk of incident AF in the long run. Greater increases in hsTnT and NT-proBNP were associated with a higher risk of AF.^47, 48^ Additionally, adding the change in NT-proBNP concentration into an established AF risk prediction model, CHARGE-AF, can modestly improve the prediction ability.^47, 49^ Our study findings have added evidence that a weight loss-focused ILI could be a substantial primary intervention approach in preventing AF onset in obese and overweight populations. The effectiveness of the intervention could be quantified by monitoring biomarkers’ levels that are AF pathway-related at both clinical and population levels.

Our study showed some strengths. First, the PREDIMED-Plus trial is a large randomized study to test the effect of an intensive lifestyle intervention focusing on weight loss for the primary prevention of CVD in the context of an erMedDiet. Second, our trial repeatedly measured biomarkers related to AF pathways through the follow-up years. Biomarkers changes were more informative than a single-time measurement to evaluate the ILI effect. They also allowed us to investigate how changes in biomarkers might affect AF development, and whether different components of the intervention mediated these effects. Third, our trial had a relatively large sample size, excellent retention of the study participants, and great reproducibility.

## Study limitations

There were limitations in this study. First, in the PREDIMED-Plus trial, older individuals who were overweight or obese and primarily of European ancestry were selected, and those with CVD were excluded. The study population limited the generalizability of our findings to the general population and other low-risk populations of CVD. Second, the ILI implemented in the trial was weight loss focused, which might mask the direct effects of physical activity and the erMedDiet on biomarkers changes while the effects through weight loss were dominating. Third, in the mediation analysis, we assumed an interaction between the exposure and the mediator. There could have been more complex interactions among the three ILI components. Moreover, the other two ILI components might introduce confounding to the mediator-outcome relationship affected by the ILI. As such, physical activity and erMedDiet could be confounders of the relationship between weight loss and hsCRP change over five years which was affected by the ILI). This mediator-outcome confounding would violate one of the assumptions for estimating natural direct or indirect effects and requires g-methods to address the issue.

## Conclusions

Weight loss-focused lifestyle intervention was beneficially associated with hsCRP, NT-proBNP, and 3-NT changes over time, highlighting mechanisms and pathways explaining how lifestyles may affect atrial substrate and, hither trough, AF development.

## Data Availability

All data produced in the present study are available upon reasonable request to the authors.

## Disclosure

None.

## Funding

Research reported in this publication was supported by the National Heart, Lung, And Blood Institute of the National Institutes of Health under Award Number R01HL137338. The content is solely the responsibility of the authors and does not necessarily represent the official views of the National Institutes of Health.

The PREDIMED-Plus trial was supported by the official funding agency for biomedical research of the Spanish government, ISCIII, through the Fondo de Investigación para la Salud (FIS), which is co-funded by the European Regional Development Fund (PI13/00673, PI13/00492, PI13/00272, PI13/01123, PI13/00462, PI13/00233, PI13/02184, PI13/00728, PI13/01090, PI13/01056, PI14/01722, PI14/0147, PI14/00636, PI14/00972, PI14/00618, PI14/00696, PI14/01206, PI14/01919, PI14/00853, PI14/01374, PI16/00473, PI16/00662, PI16/01873, PI16/01094, PI16/00501, PI16/00533, PI16/00381, PI16/00366, PI16/01522, PI16/01120, PI17/00764, PI17/01183, PI17/00855, PI17/01347, PI17/00525, PI17/01827, PI17/00532, PI17/00215, PI17/01441, PI17/00508, PI17/01732, PI17/00926, PI19/00957, PI19/00386, PI19/00309, PI19/01032, PI19/00576, PI19/00017, PI19/01226, PI19/00781, PI19/01560, PI19/01332), the European Research Council Advanced Research Grant 2013–2018 (340918), the Recercaixa grant 2013ACUP00194, grants from the Consejería de Salud de la Junta de Andalucía (PI0458/2013; PS0358/2016, PI0137/2018), the PROMETEO/2017/017 grant from the Generalitat Valenciana, the SEMERGEN grant and FEDER funds (CB06/03).

## Data sharing

Data collaboration for PREDIMED-Plus study is guided by the Data Sharing and Management guide. We follow a controlled data collaboration model, using anonymised (de-identified) study data only, for collaborating with approved researchers. Requests are considered by the PREDIMED-Plus Steering Committee. Please see https://www.predimedplus.com/en/project/#top for detail.

---

**Supplemental Table 1.** Effect of an intensive lifestyle intervention (ILI) on selected circulating biomarkers stratified by sex and age, PREDIMED-Plus trial.

|  |  | Difference ILI - control |  |  |
| --- | --- | --- | --- | --- |
|  |  | Y3 vs baseline | Y5 vs baseline | p-value for interaction |
| Male (n=278) | PICP | -2% (-12%, 9%) | -3% (-13%, 8%) |  |
|  | Hs-TnT | -4% (-11%, 3%) | -3% (-9%, 4%) |  |
|  | HsCRP | -15% (-33%, 7%) | -27% (-42%, -8%) |  |
|  | 3-NT | -13% (-26%, 3%) | -8% (-22%, 8%) |  |
|  | NT-proBNP | -7% (-22%, 11%) | -19% (-33%, -3%) |  |
| Female (n=193) | PICP | 1% (-12%, 16%) | 4% (-10%, 19%) | 0.65 |
|  | Hs-TnT | 1% (-7%, 8%) | -4% (-11%, 4%) | 0.42 |
|  | HsCRP | -4% (-22%, 18%) | 5% (-15%, 30%) | 0.31 |
|  | 3-NT | -6% (-21%, 13%) | -25% (-38%, -10%) | 0.15 |
|  | NT-proBNP | -2% (-23%, 25%) | -6% (-27%, 20%) | 0.63 |
| Age <65 (n=209) | PICP | 0% (-10%, 11%) | -3% (-13%, 8%) |  |
|  | Hs-TnT | -4% (-11%, 4%) | -2% (-9%, 7%) |  |
|  | HsCRP | -12% (-31%, 13%) | -22% (-39%, 1%) |  |
|  | 3-NT | -8% (-25%, 12%) | -8% (-25%, 13%) |  |
|  | NT-proBNP | -14% (-31%, 8%) | -14% (-32%, 7%) |  |
| Age ≥65 (262) | PICP | -1% (-13%, 12%) | 3% (-10%, 17%) | 0.81 |
|  | Hs-TnT | 0% (-7%, 6%) | -4% (-10%, 3%) | 0.30 |
|  | HsCRP | -11% (-28%, 11%) | -11% (-28%, 11%) | 0.89 |
|  | 3-NT | -11% (-24%, 3%) | -21% (-32%, -8%) | 0.09 |
|  | NT-proBNP | 5% (-13%, 27%) | -12% (-27%, 6%) | 0.46 |

Notes: Mixed models were used. Results are shown as estimate (95% confidence interval). All the concentrations of biomarkers were log-transformed. Percent changes in concentrations of biomarkers comparing intervention and control arm were presented, calculated as $[\exp(\text{coefficient}) - 1] \times 100$ . Follow-up exams occurred in years 3 and 5. PICP: C-terminal propeptide of procollagen type I; hsTnT: high sensitivity troponin T; hsCRP: high-sensitivity C reactive protein; 3-NT: 3-nitrotyrosine; NT-proBNP: N-terminal propeptide of B-type natriuretic peptide.

**Supplemental Table 2.** Changes in log-transformed concentrations of biomarkers through 3 and 5 years according to the intervention group and follow-up time, missing value imputed by multiple imputation by chained equation (MICE), PREDIMED-Plus trial.

|  | Difference ILI - control |  |
| --- | --- | --- |
|  | Y3 vs. baseline | Y5 vs. baseline |
| PICP | 1% (-8%, 11%) | 1% (-8%, 11%) |
| HsTnT | -2% (-7%, 4%) | -3% (-9%, 4%) |
| hsCRP | -7% (-22%, 11%) | -12% (-26%, 5%) |
| 3-NT | -10% (-22%, 4%) | -13% (-24%, 1%) |
| NT-proBNP | -3% (-18%, 13%) | -14% (-27%, 1%) |

Notes: Mixed models were used. Results are shown as estimate (95% confidence interval). All the concentrations of biomarkers were log-transformed. Percent changes in concentrations of biomarkers comparing intervention and control arm were presented, calculated as $[\exp(\text{coefficient}) - 1] \times 100$ . PICP: C-terminal propeptide of procollagen type I; hsTnT: high sensitivity troponin T; hsCRP: high-sensitivity C reactive protein; 3-NT: 3-nitrotyrosine; NT-proBNP: N-terminal propeptide of B-type natriuretic peptide.

**Supplemental Table 3.** Changes in log-transformed concentrations of biomarkers through 3 and 5 years according to the intervention group and follow-up time, adjust for covariates at baseline, PREDIMED-Plus trial.

|  | Difference ILI - control |  |
| --- | --- | --- |
|  | Y3 vs. baseline | Y5 vs. baseline |
| PICP | -1% (-9%, 9%) | 1% (-8%, 10%) |
| HsTnT | -2% (-7%, 3%) | -3% (-8%, 3%) |
| hsCRP | -12% (-26%, 3%) | -16% (-29%, -1%) |
| 3-NT | -8% (-19%, 5%) | -13% (-23%, -1%) |
| NT-proBNP | -7% (-19%, 8%) | -15% (-27%, -2%) |

Notes: Mixed models were used, adjusting for sex, age, education level, marital status, smoking, Mediterranean diet adherence score, and total physical activity at baseline. Results are shown as estimate (95% confidence interval). All the concentrations of biomarkers were log-transformed. Percent changes in concentrations of biomarkers comparing intervention and control arm were presented, calculated as $[\exp(\text{coefficient}) - 1] \times 100$ . Follow-up exams occurred in years 3 and 5. PICP: C-terminal propeptide of procollagen type I; hsTnT: high sensitivity troponin T; hsCRP: high-sensitivity C reactive protein; 3-NT: 3-nitrotyrosine; NT-proBNP: N-terminal propeptide of B-type natriuretic peptide.

**Supplemental Table 4.** Changes in log-transformed concentrations of biomarkers through 3 and 5 years according to the intervention group and follow-up time, adjust for eGFR, PREDIMED-Plus trial.

|  | Difference ILI - control |  |
| --- | --- | --- |
|  | Y3 vs baseline | Y5 vs baseline |
| PICP, mg/mL | -1% (-9%, 8%) | 0% (-8%, 9%) |
| Hs-TnT, ng/L | -2% (-7%, 3%) | -3% (-8%, 2%) |
| hsCRP, mg/dL | -11% (-25%, 4%) | -15% (-28%, 0%) |
| 3-NT, nM | -10% (-21%, 1%) | -16% (-25%, -4%) |
| NT-proBNP, pg/mL | -5% (-17%, 10%) | -13% (-25%, 1%) |

Notes: Mixed models were used. Results are shown as estimate (95% confidence interval). All the concentrations of biomarkers were log-transformed. Percent changes in concentrations of biomarkers comparing intervention and control arm were presented, calculated as $[\exp(\text{coefficient}) - 1] \times 100$ . Follow-up exams occurred in years 3 and 5. PICP: C-terminal propeptide of procollagen type I; hsTnT: high sensitivity troponin T; hsCRP: high-sensitivity C reactive protein; 3-NT: 3-nitrotyrosine; NT-proBNP: N-terminal propeptide of B-type natriuretic peptide.

## Notes

### Competing Interest Statement

The authors have declared no competing interest.

### Clinical Trial

The PREDIMED--Plus trial was registered at the International Standard Randomized Controlled Trial (ISRCT; http://www.isrctn.com/ISRCTN89898870) with number 89898870 and a registration date of 24 July 2014.

### Clinical Protocols

https://www.predimedplus.com/wp-content/uploads/2018/11/Protocolo-PREDIMED-Plus_Eng.pdf

### Author Declarations

Ethics Committee of Clinical Research of the Department of Health of the Government of Navarra, Research Ethics Committee of the Balearic Islands, and Basque Country Clinical Research Ethics Committee gave ethical approval for this work. The institutional review boards of all the recruiting centers approved the study protocol.

### Summary of Updates

Section Methods-Statistical Analysis, Tables 2&3, Supplemental Tables 1-4 notes updated to clarify results presentation and statistical significance.

